# The unified proposal for classification of human respiratory syncytial virus below the subgroup level

**DOI:** 10.1101/2024.02.13.24302237

**Authors:** Stephanie Goya, Christopher Ruis, Richard A. Neher, Adam Meijer, Ammar Aziz, Angie S. Hinrichs, Anne von Gottberg, Cornelius Roemer, Daniel G. Amoako, Dolores Acuña, Jakob McBroome, James R. Otieno, Jinal N. Bhiman, Josie Everatt, Juan C. Muñoz-Escalante, Kaat Ramaekers, Kate Duggan, Lance D. Presser, Laura Urbanska, Marietjie Venter, Nicole Wolter, Teresa C. T. Peret, Vahid Salimi, Varsha Potdar, Vítor Borges, Mariana Viegas

## Abstract

A globally implemented unified classification for human respiratory syncytial virus (HRSV) below the subgroup level remains elusive. Here, we formulate the global consensus of HRSV classification based on the challenges and limitations of our previous proposals and the future of genomic surveillance. From a high-quality dataset of 1,480 HRSV-A and 1,385 HRSV-B genomes submitted to NCBI and GISAID up to March 2023, we categorized HRSV-A/B sequences into lineages based on phylogenetic clades and amino acid markers. We defined 24 lineages within HRSV-A and 16 within HRSV-B, providing guidelines for prospective lineages definition. Our classification demonstrated robustness in its applicability to both complete and partial genomes. In addition, it allowed the observation of notable lineage replacements and the identification of lineages exclusively detected since the COVID-19 pandemic. We envision that this unified HRSV classification proposal will strengthen and facilitate HRSV molecular epidemiology on a global scale.

## Introduction

Significant progress has been achieved in developing interventions against the human respiratory syncytial virus (HRSV), a leading cause of acute lower respiratory tract infection in children, elderly, and immunocompromised individuals. In 2023, the U.S. Food and Drug Administration (FDA) and the European Medicines Agency (EMA) approved the first HRSV vaccines (1,2). Simultaneously, a monoclonal antibody was approved for widespread use in infants and not limited to high-risk and premature children (3). The availability of HRSV immunization highlights the role of molecular epidemiology as a tool to monitor their efficacy. Nevertheless, a standardized HRSV genotype system has yet to be defined and implemented.

In 2022, HRSV was designated as “*Orthopneumovirus hominis*” species within the *Pneumoviridae* family. Below species level, there are two antigenic groups known as HRSV subgroup A (HRSV-A) and B (HRSV-B), that were previously referred to as subtypes (4–6). Within each subgroup, genotypes were initially defined based on statistically supported phylogenetic clades inferred with the second hypervariable region (2HR) of the G gene (Figure 1A-B) (7). The G gene, encoding the attachment glycoprotein, exhibits the highest genetic and antigenic variability. Notably, this gene has undergone a duplication of a 72-nt fragment in HRSV-A and 60-nt in HRSV-B (Figure 1B) (8,9).

**Figure 1.**
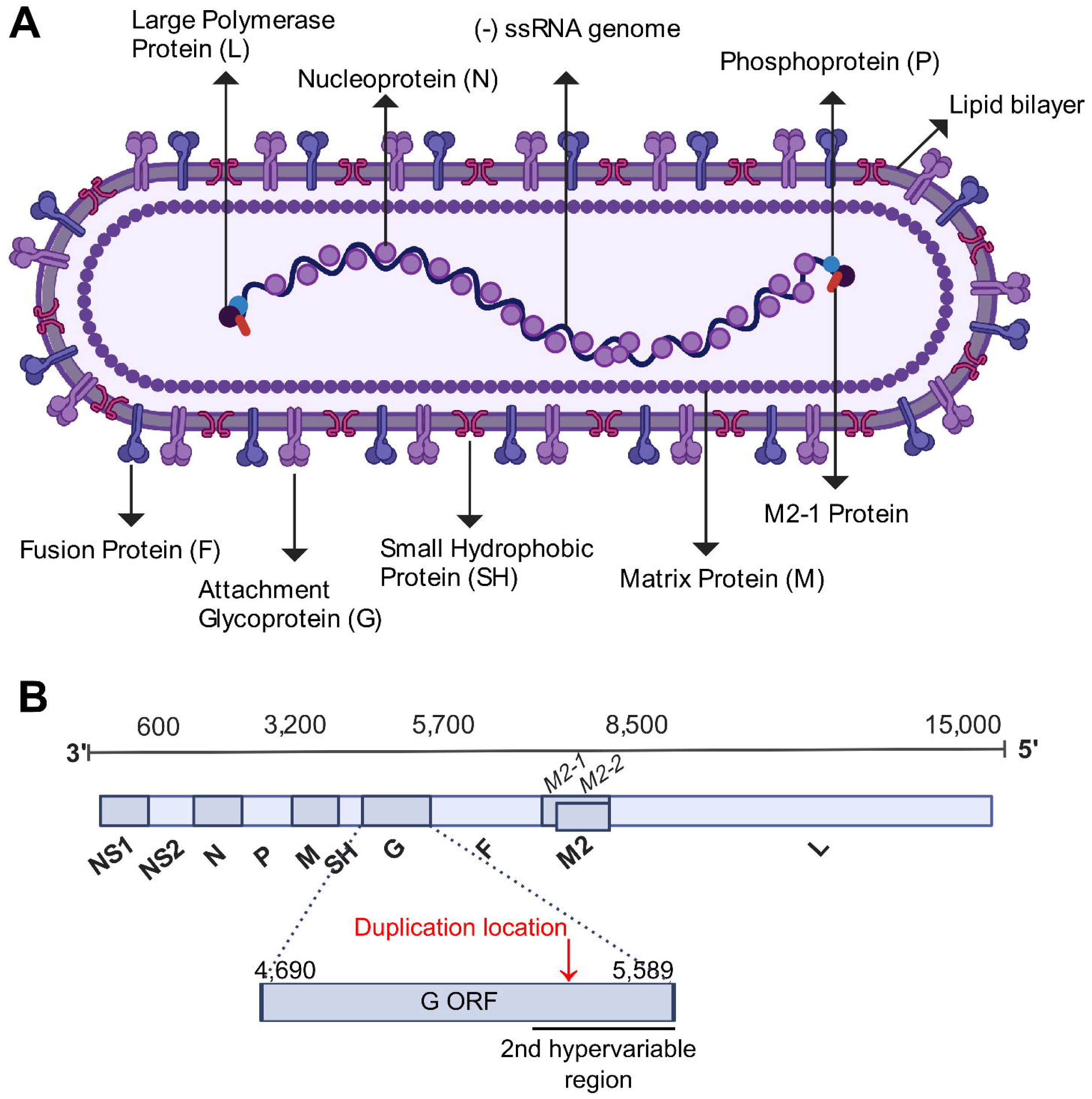
The structure and genome of HRSV. (A) Schematic of the HRSV virion structure detailing the location of structural proteins, created with BioRender.com. (B) Schematic of the HRSV genome organization with the approximated location of genes highlighted (the exact location slightly differs between subgroups and strains). The location of the second hypervariable region in the G gene, used originally for molecular epidemiology classification, is detailed. The location of the G gene 72-nt duplication in HRSV-A and 60-nt duplication in HRSV-B occurs is denoted with a red arrow.

To identify emerging genotypes, genetic distances between phylogenetic clades and distinctive genetic features were used, accompanied by variable nomenclature based on the gene (GA1–GA7 in HRSV-A and GB1–GB4 in HRSV-B), country and subgroup (SAB1–SAB4 for South African genotypes in HRSV-B) or city/province (NA1-NA2 [Niigata], ON1 [Ontario] in HRSV-A and BA1-BA9 [Buenos Aires] in HRSV-B) (7–15). Since 2019 alternative phylogenetic reclassifications were proposed using G gene sequences or complete genomes. From one side, the use of the G gene allows laboratories without capacity for whole genome sequencing (WGS) to perform molecular epidemiology studies (16,17). From the other side, complete genomes allow the surveillance of all viral genes, including the F gene encoding the fusion protein, which is the target of monoclonal antibodies, and the basis of approved vaccines against HRSV (Figure 1A) (18,19).

The milestones in HRSV interventions have renewed interests to address the challenge of classifying HRSV below the subgroup level. This prompted establishment of the HRSV Genotyping Consensus Consortium (RGCC), formed by HRSV and virus evolution experts aiming to standardize the HRSV classification system. Here, we present a framework for HRSV classification below the subgroup level, based on current knowledge of HRSV diversity and evolution, focused on practical implementation for molecular epidemiology.

## Methodology

### HRSV sequences dataset

The HRSV complete genomes available from NCBI GenBank (https://www.ncbi.nlm.nih.gov/labs/virus) and GISAID EpiRSV (https://gisaid.org/) up to March 11, 2023 were downloaded using a filter for sequence length above 14,000-nt, obtained from human hosts and including the year and country of the sample collection. Sequences were categorized into files based on their reported subgroup resulting in 2,744 HRSV-A and 2,443 HRSV-B genomes (Supplementary Figure 1). Sequences containing nucleotide ambiguities, indicative of inadequate sequencing depth, were reserved for epidemiological analysis but excluded from formal lineage definition. Removal of genomes with any nucleotide ambiguities was performed using BBmap (reformat, version Jan-2021). To ensure diversity without redundancy, BBmap (dedupe, version Feb-2020) was used to remove identical sequences, preserving one representative.

Sequences were aligned with MAFFT v7.490, and alignments were inspected and corrected with Aliview v1.28, mainly in the G gene (20,21). Furthermore, alignment ends were trimmed to encompass complete genomes from the first codon of the first gene (NS1) to the last codon of the last gene (L). Partial genomes were considered if the lack of sequence was within 50-nt of the genome ends. Genomes with nucleotide insertions or deletions causing frameshift in any open reading frame(s) were identified with RSVsurver and removed (22). Following alignment trimming, the presence of identical sequences with nucleotide differences in the trimmed region prompted another round of redundancy removal using BBmap tool, resulting in the final set of 1,538 HRSV-A and 1,387 HRSV-B genomes (Supplementary Figure 1).

### Phylogenetic analysis

Maximum likelihood phylogenetic trees were constructed with IQ-TREE v2.2.0 (details in Supplementary Material). Monophyletic clades were considered statistically supported when SH-aLRT value was ≥80% and UFBoot2 value was ≥90% (23,24). Phylogenetic trees were visualized with Figtree v1.4.4 and Auspice. Temporal signal was assessed with TempEst v1.5.3, and molecular-clock phylogenies were inferred with TreeTime (25).

The ancestral sequence reconstruction was inferred using Augur bioinformatic toolkit v23.1.0 (26). Recombination events were assessed with alignment-based method using RDP4 and phylogenetic-based TreeKnit (Supplementary Material).

The amino acid substitutions linked to the clades in the tree were inferred using Augur and the initial screening of lineages was automated with Autolin (27). Amino acid comparison among monophyletic clusters underwent manual curation to rectify conflicts arising from internal (nested) lineages and the confirmation of the lineage-defining amino acids in more than 90% of the clade’s sequences. Results from this study are available: https://github.com/rsv-lineages/Classification_proposal.

## Results and Discussion

### Baseline agreements on the HRSV classification definition

The proposed classification establishes “HRSV lineages” for viruses below subgroup level. Recent studies have demonstrated superior resolution provided by complete genomes in constructing HRSV phylogenetic trees (16,18,19). Anticipating improvement of WGS, we defined a classification system based on maximum likelihood phylogenetic trees inferred from complete HRSV genomes. The maximum likelihood algorithm formulates hypotheses about the evolutionary relationships among sequences, and the implementation within IQ-TREE dealing with large datasets makes it particularly well-suited to assert HRSV genomic phylogeny including sequences collected >50 years ago (23).

The definition of complete genome used in this study spans from the first codon of the first gene (NS1) to the last codon of the last gene (L). “Almost” complete genomes were considered if the sequence information gaps were within a 50-nt window at the genome ends. For a robust genetic dataset, only genomes without nucleotide ambiguities (according to the IUPAC code for nucleotide degeneracy) were used to define lineages.

### Composition of the genomic dataset used for lineage classification

Applying the established baseline agreements, we gathered 1,538 HRSV-A and 1,387 HRSV-B high-quality, public genomes. Analysis of the genomic dataset revealed a limited global HRSV genomic surveillance until 2007, with <20 genomes deposited annually (Figure 2A, Supplementary Figure 2). Since 2008, there was improvement in the number of genomes and representation of countries, probably attributed to increased adoption of sequencing technology. Additionally, a significant surge occurred post-2021, probably driven by expansion of viral genomics since the SARS-CoV-2 pandemic and the approval of the HRSV prophylactic treatments (Figure 2A, Supplementary Figure 2). Considering delays in genome deposition in public databases, the number of genomes in 2022 may rise if the submission dates to the genomic databases postdate the data used in this study. Regarding geographic representation, nine countries (Australia, United Kingdom, New Zealand, USA, Argentina, Kenya, Morocco, Netherlands, and Brazil) submitted >100 genomes, only United Kingdom achieved uninterrupted surveillance since 2008, but Australia deposited the most genomes globally (Figure 2B).

**Figure 2.**
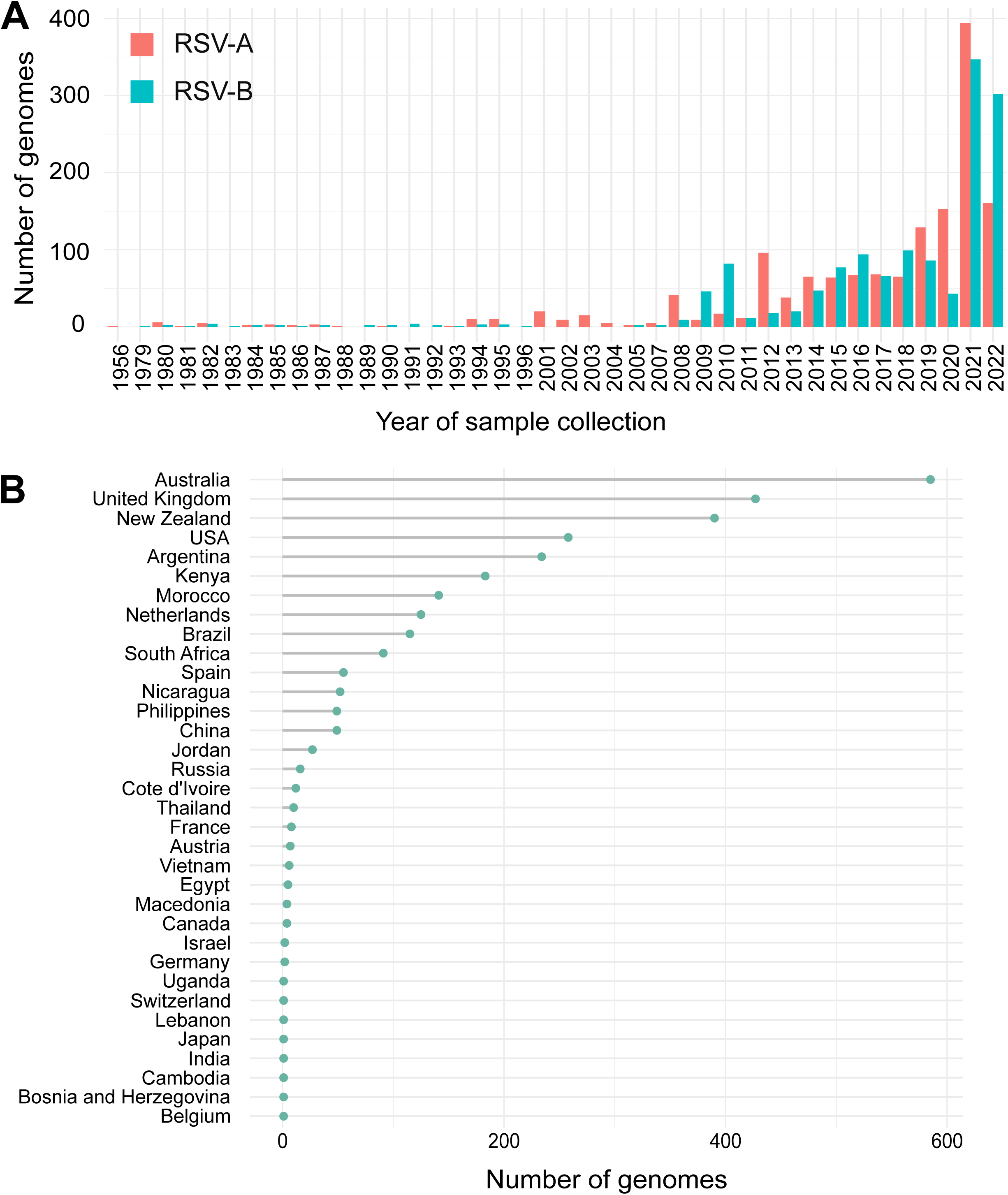
The current HRSV genomics surveillance landscape. Number of HRSV genomes meeting the inclusion criteria used for the classification (NCBI Virus and GISAID review up to March 11, 2023) according to the year of sample collection and subgroup (A) and their country of sample collection (B).

### Defining the root and the final dataset of HRSV phylogenetic trees

We reconstructed separate maximum likelihood phylogenetic trees for the HRSV-A/B datasets. The first step toward a reliable classification system requires precise determination of the phylogenetic tree root. Two approaches were used: 1) utilization of an outgroup, a conventional method for inferring the tree root using sequences known to be evolutionarily distant, and 2) phylodynamic analysis, integrating temporal and phylogenetic patterns in virus evolution (Supplementary Material, Supplementary Figure 3). Both approaches consistently identified the same root for each subgroup cluster. Phylodynamic analysis also identified 58 outlier sequences for HRSV-A and two for HRSV-B that were excluded from lineage designation. The final dataset considered for lineage designation comprised 1,480 HRSV-A and 1,385 HRSV-B genomes (Supplementary Table 1 and 2).

### The HRSV lineage definition

An HRSV lineage was defined as a statistically supported monophyletic cluster comprising ≥10 sequences and characterized by ≥5 amino acid substitutions compared to the parental lineage. The lineage-defining amino acids, present in ≥90% of the sequences within the clade, may be found in any of the viral proteins.

Phylogenetic classifications vary among viral species, and all try to ensure the defined clustering reflects the heterogeneity of the viral population, therefore requiring the consideration of unique evolutionary characteristics of each viral species, the use of arbitrary thresholds and the feasibility for long-term use (28–30). There is inherent bias in any classification due to sequence availability and spatiotemporal representation of viral diversity. Therefore, HRSV lineage definition does not include criteria of sequences from different outbreaks or countries to ensure early detection of new lineages.

We observed that presence of distinctive amino acids shared by sequences of a phylogenetic clade in comparison to the parental lineage is a simple method to identify a new lineage. Methods (i.e. average nucleotide genetic distances, average patristic distances, or patristic distances between nodes) need phylogenies with complete datasets to define new categories, becoming complex with rapid increases of available sequences (16–19). In this proposal, different amino acid thresholds were initially screened in an automated manner, ranging from 1-10 lineage-defining amino acids (Supplementary Material). The number of small lineages decreases as the number of lineage-defining amino acids increases, and five amino acids resulted in an intermediate complexity of lineages defined for both HRSV subgroups. We acknowledge that other thresholds could be useful, but we emphasize that the key to establishing a global consensus is clear operational guidelines and a robust classification, two aspects that our proposal fulfills.

### a) Definition of the lineage’s nomenclature

Lineage nomenclature integrates the HRSV subgroup letter and ascending ordinal numbers, separated by dots to represent nested lineages (Figure 3A-B). Furthermore, a distinct nomenclature was assigned to the 72-nt (24 amino acids) and 60-nt (20 amino acids) G-gene duplication within HRSV-A/B, respectively. This genetic event is epidemiologically relevant, as viruses with this duplication have exclusively been detected since 2016 (31–34). To facilitate tracking these viruses, we use the alias “D”, specifically A.D (historically, ON1 genotype) and B.D (historically, BA genotype) for HRSV-A/B respectively, and nested lineages with increasing ordinal numbers.

**Figure 3.**
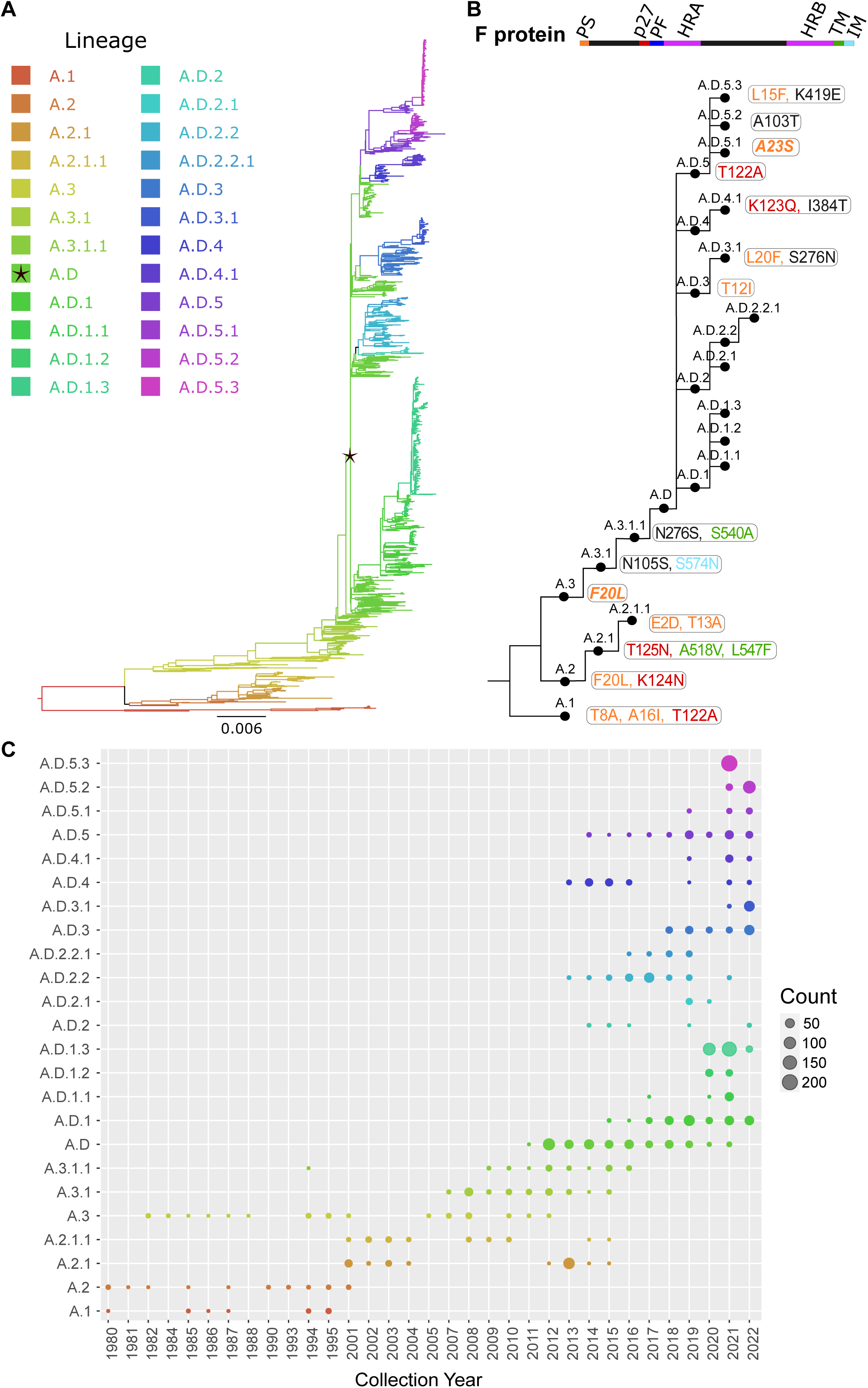
HRSV-A lineages classification and seasonality. (A) The HRSV-A maximum likelihood phylogenetic tree (1,480 sequences) colored according to lineage classification is shown. The A.D lineage defined by the 72-nt duplication in the G gene is denoted with a black star. Scale bar indicates substitutions per site. (B) Simplified scheme of the lineage designation to highlight the presence of nested lineages. The amino acid changes in the F protein are listed next to lineage name and colored according to their location in the fusion protein. (C) Temporal distribution of the HRSV-A lineages including all the genomes available in public databases up to March 2023 (2,744 sequences).

In summary, letters A and B indicate the HRSV subgroup at the beginning of the lineage name, C is unused, and D serves as an alias for 72-nt and 60-nt duplication within the G gene. In addition, aliases starting from E are limited to three numerical levels of nested lineages, preventing indefinite accumulation of numbers. For example, B.D.4.1.1 lineage has descendant lineages named B.D.E.1 - B.D.E.4 instead of B.D.4.1.1.1 - B.D.4.1.1.4, where “E” represents “4.1.1” (Figure 3A-B). The nomenclature is based on the tree topology, reflecting the order of the nodes from the root to the tips, but it is unrelated to the sequence collection date or date of the most recent common ancestor of the lineage.

To remain functional, a nomenclature system requires periodic updates as new lineages emerge. Therefore, we have established two open repositories on GitHub containing definitions of each lineage, signature mutations, and representative sequences. These repositories, available at https://github.com/HRSV-lineages/lineage-designation-A and https://github.com/HRSV-lineages/lineage-designation-B are intended to provide up-to-date definitions, and serve as a platform for discussion and designation of novel lineages.

### b) Identification of lineages within the HRSV-A and HRSV-B rooted trees

We reconstruct ancestral sequences at the root of the phylogenetic trees. While these sequences are not biologically real, they serve as surrogate parental lineages during initial classification. Identifying monophyletic clusters with ≥10 sequences and ≥5 amino acid changes compared to the reconstructed root sequence, we defined three HRSV-A lineages (A.1 – A.3) and four HRSV-B lineages (B.1 – B.4). Two sequences, (EPI-ISL-15771600_USA_1956 and MG642074_USA_1980) could not be classified perhaps because they belong to underrepresented extinct lineages.

The first lineages underwent further analysis to identify nested lineages in an iterative manner, resulting in a total of 24 lineages within HRSV-A, and 16 within HRSV-B (Figure 3 and Figure 4). Close to the root of the HRSV-B tree, extinct lineages were underrepresented, comprising <10 sequences but featuring >5 distinct amino acids (B.1, B.3, B.4). Despite the low number of sequences, we included them as lineages to trace evolutionary branches that gave rise to currently circulating lineages. In addition, A.D.2 is slightly below the sequences threshold, but we kept the lineage category to emphasize the common ancestor among A.D.2.1 and A.D.2.2.

**Figure 4.**
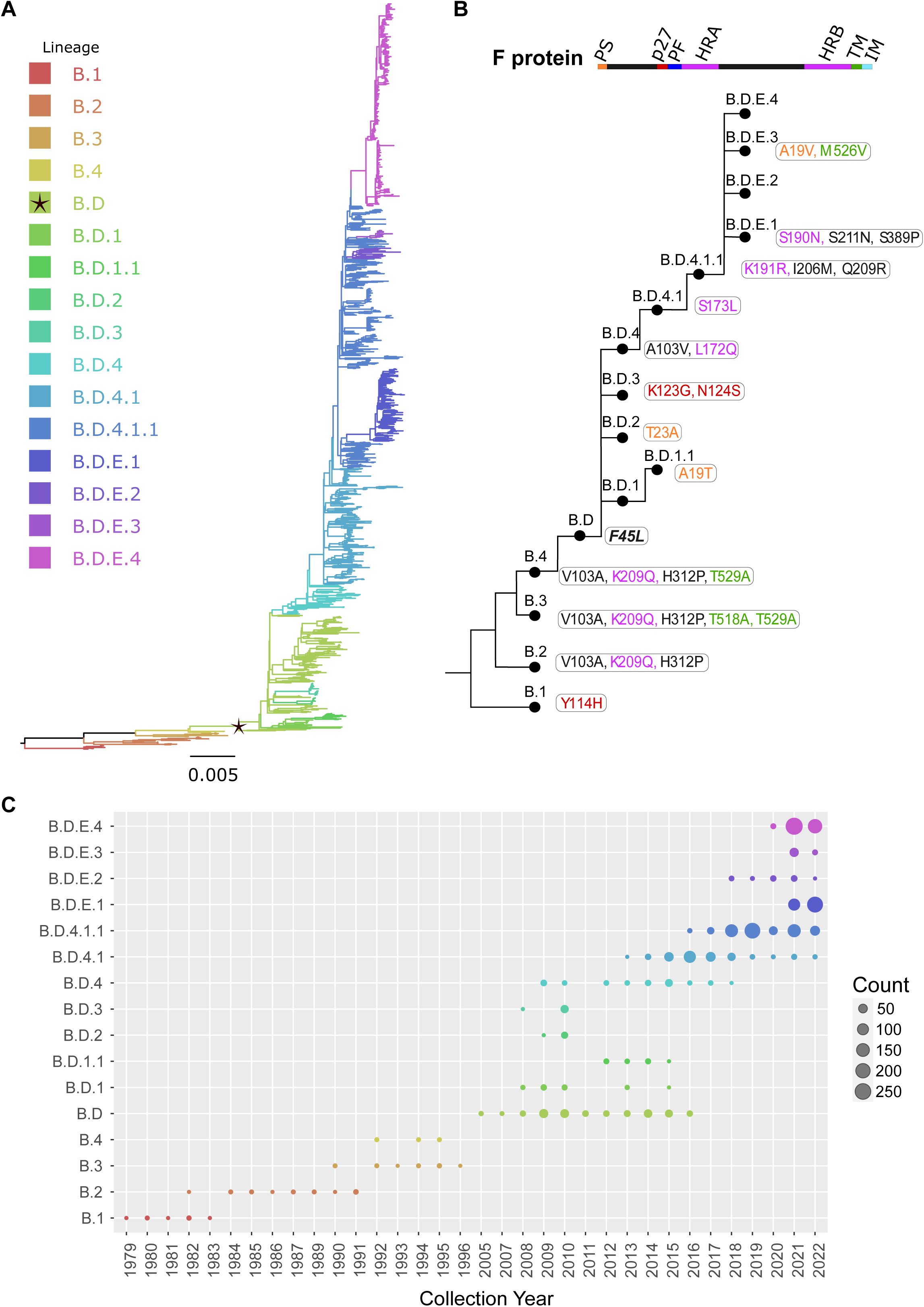
HRSV-B lineages classification and seasonality. (A) The HRSV-B maximum likelihood phylogenetic tree (1,385 sequences) colored according to lineage classification is shown. The B.D lineage defined by the 60-nt duplication in the G gene is denoted with a black star. Scale bar indicates substitutions per site. (B) Simplified scheme of the lineage designation to highlight the presence of nested lineages. The amino acid changes in the F protein are listed next to lineage name and colored according to their location in the fusion protein. (C) Temporal distribution of the HRSV-B lineages including all the genomes available in public databases up to March 2023 (2,443 sequences).

We scrutinized the presence and absence of the duplication in the G gene across each tree. While patterns were mostly as expected with a single historical duplication event, some genomes within the clade with the duplication in G, lacked the duplication. The dispersed association of these sequences in the phylogenetic tree, rather than forming a monophyletic cluster, suggests the virus did not lose the nucleotide duplication (Supplementary Figure 4). Instead, similar read length to the duplication region of certain short-read NGS technologies potentially masks the presence of the duplication when used in the consensus genome assembly with reference sequences that do not possess the nucleotide duplication. Therefore, we recommend the use of NGS data with quality filtered reads with a length greater than 150-nt to avoid this problem.

Lineage-defining amino acids were present in all HRSV proteins, the majority identified within the G protein (Tables 1 and 2). Also, the polymerase L protein was noteworthy, contributing to the distinction of 21 out of 24 HRSV-A lineages and 15 out of 16 HRSV-B lineages (Table 1 and Table 2). Interestingly, the F protein contributed to define 14 lineages in HRSV-A and 13 in HRSV-B (Figure 3B, Figure 4B). Both the G and F surface glycoproteins are under selection pressure from humoral immunity and exhibit a robust phylogenetic signal (18,32). While the G protein displays substantial nucleotide and amino acid sequence plasticity, the F protein experiences strong negative selection, likely attributed to functional/structural constraints (35). In fact, the fusion peptide is the only region in F without lineage-defining amino acids (Figure 3B, Figure 4B). While the low diversity of the F protein is promising for HRSV interventions, monitoring the F protein during their global implementation is essential.

**Table 1.**
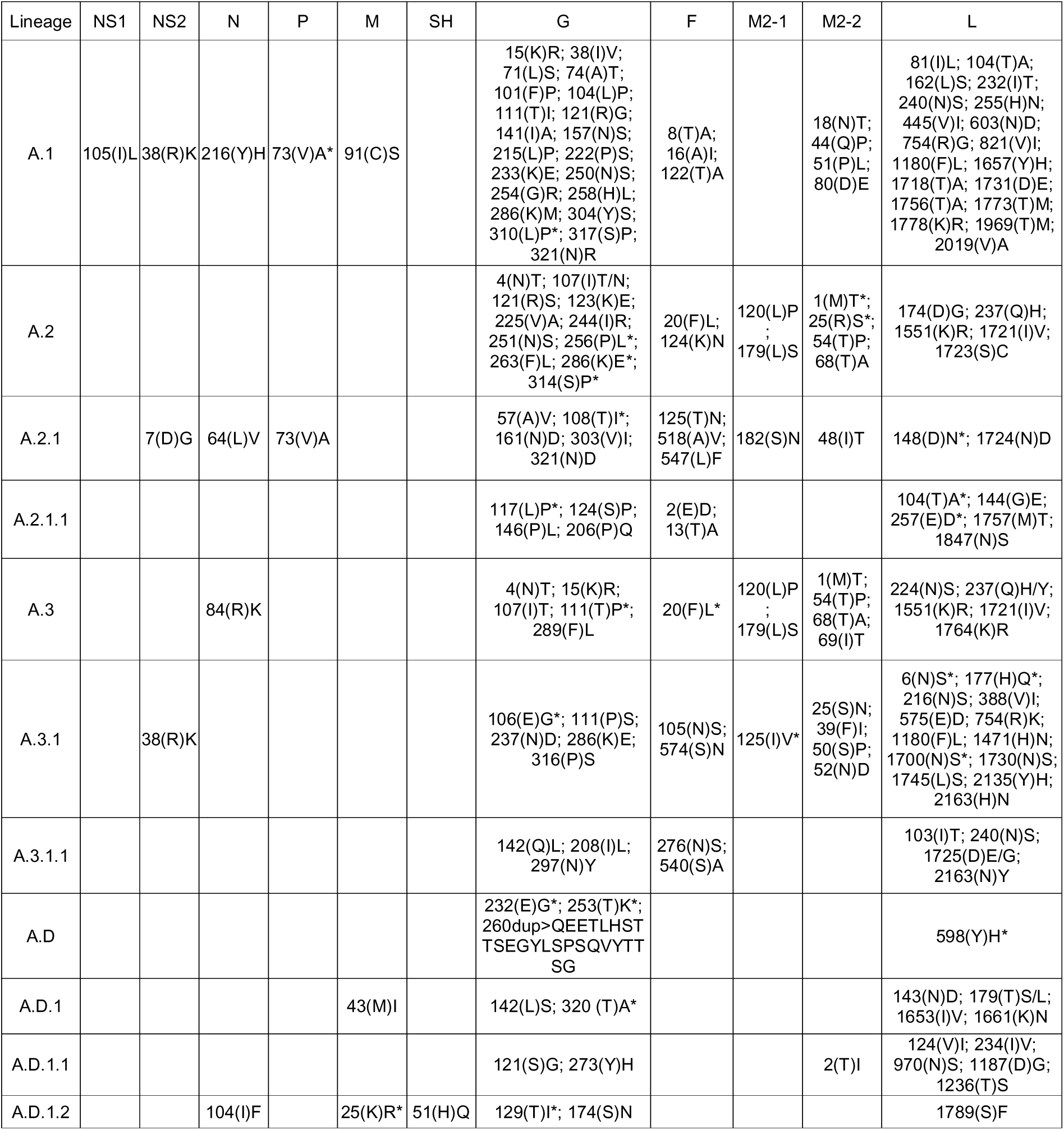
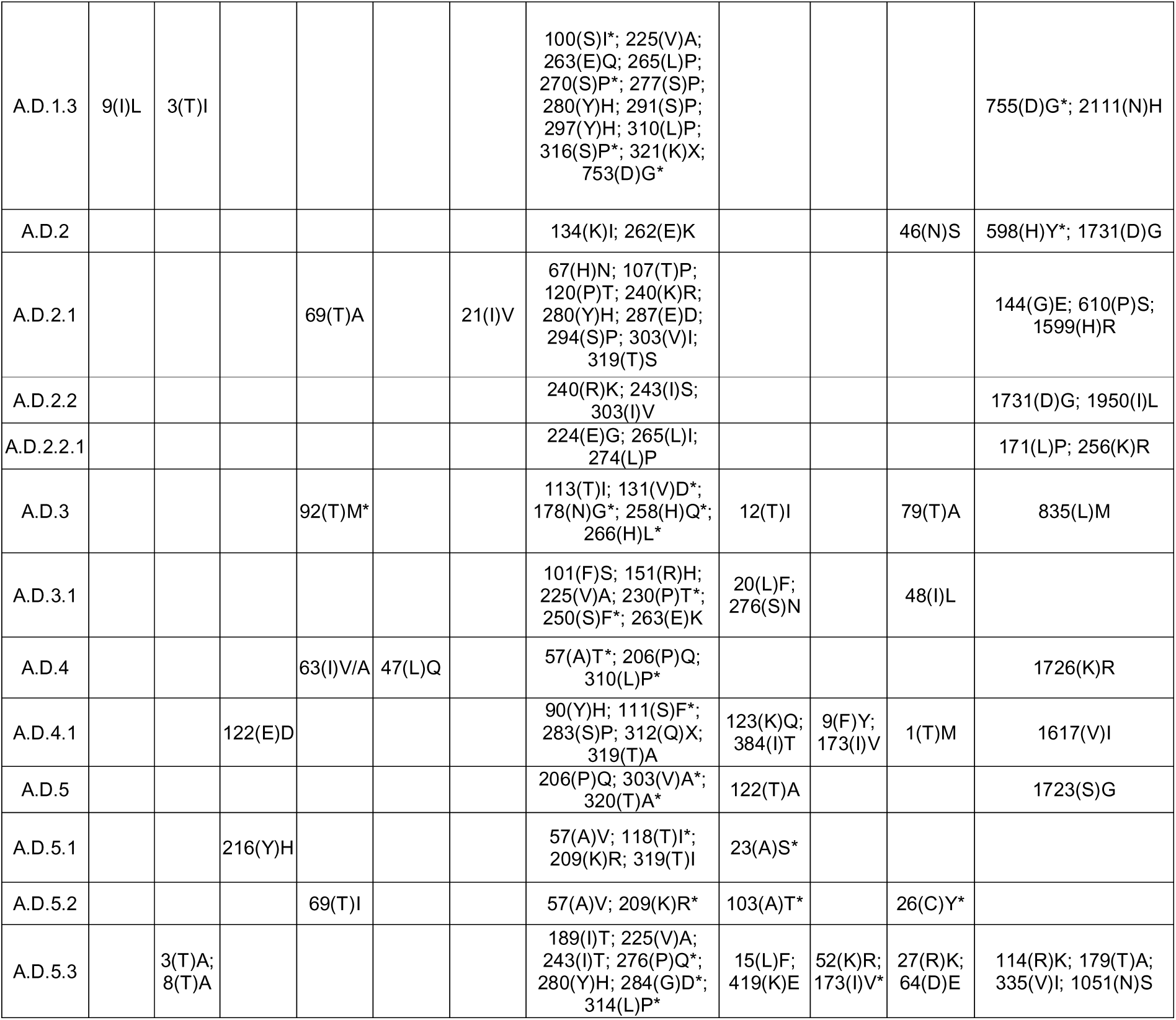
Amino acids defining lineages in HRSV-A. The lineage-defining amino acids within each viral protein are detailed. This description provides the location of the amino acid using the genome coordinates hRSV/A/England/397/2017 (lineage A.D.2.2.1, NCBI Virus accession number: PP109421.1, GISAID EpiRSV accession number: EPI_ISL_412866), followed by the amino acid found in the parental lineage in parentheses. Finally, the amino acid substitution that drives it to become a lineage is specified. Amino acid changes detected in not all but ≥90% of the lineage sequences are indicated with *. The letter X is used to represent an amino acid deletion.

| Lineage | NS1 | NS2 | N | P | M | SH | G | F | M2-1 | M2-2 | L |
| --- | --- | --- | --- | --- | --- | --- | --- | --- | --- | --- | --- |
| A.1 | 105(I)L | 38(R)K | 216(Y)H | 73(V)A* | 91(C)S |  | 15(K)R; 38(I)V;<br>71(L)S; 74(A)T;<br>101(F)P; 104(L)P;<br>111(T)I; 121(R)G;<br>141(I)A; 157(N)S;<br>215(L)P; 222(P)S;<br>233(K)E; 250(N)S;<br>254(G)R; 258(H)L;<br>286(K)M; 304(Y)S;<br>310(L)P*; 317(S)P;<br>321(N)R | 8(T)A;<br>16(A)I;<br>122(T)A |  | 18(N)T;<br>44(Q)P;<br>51(P)L;<br>80(D)E | 81(I)L; 104(T)A;<br>162(L)S; 232(I)T;<br>240(N)S; 255(H)N;<br>445(V)I; 603(N)D;<br>754(R)G; 821(V)I;<br>1180(F)L; 1657(Y)H;<br>1718(T)A; 1731(D)E;<br>1756(T)A; 1773(T)M;<br>1778(K)R; 1969(T)M;<br>2019(V)A |
| A.2 |  |  |  |  |  |  | 4(N)T; 107(I)T/N;<br>121(R)S; 123(K)E;<br>225(V)A; 244(I)R;<br>251(N)S; 256(P)L*;<br>263(F)L; 286(K)E*;<br>314(S)P* | 20(F)L;<br>124(K)N | 120(L)P<br>;<br>179(L)S | 1(M)T*;<br>25(R)S*;<br>54(T)P;<br>68(T)A | 174(D)G; 237(Q)H;<br>1551(K)R; 1721(I)V;<br>1723(S)C |
| A.2.1 |  | 7(D)G | 64(L)V | 73(V)A |  |  | 57(A)V; 108(T)I*;<br>161(N)D; 303(V)I;<br>321(N)D | 125(T)N;<br>518(A)V;<br>547(L)F | 182(S)N | 48(I)T | 148(D)N*; 1724(N)D |
| A.2.1.1 |  |  |  |  |  |  | 117(L)P*; 124(S)P;<br>146(P)L; 206(P)Q | 2(E)D;<br>13(T)A |  |  | 104(T)A*; 144(G)E;<br>257(E)D*; 1757(M)T;<br>1847(N)S |
| A.3 |  |  | 84(R)K |  |  |  | 4(N)T; 15(K)R;<br>107(I)T; 111(T)P*;<br>289(F)L | 20(F)L* | 120(L)P<br>;<br>179(L)S | 1(M)T;<br>54(T)P;<br>68(T)A;<br>69(I)T | 224(N)S; 237(Q)H/Y;<br>1551(K)R; 1721(I)V;<br>1764(K)R |
| A.3.1 |  | 38(R)K |  |  |  |  | 106(E)G*; 111(P)S;<br>237(N)D; 286(K)E;<br>316(P)S | 105(N)S;<br>574(S)N | 125(I)V* | 25(S)N;<br>39(F)I;<br>50(S)P;<br>52(N)D | 6(N)S*; 177(H)Q*;<br>216(N)S; 388(V)I;<br>575(E)D; 754(R)K;<br>1180(F)L; 1471(H)N;<br>1700(N)S*; 1730(N)S;<br>1745(L)S; 2135(Y)H;<br>2163(H)N |
| A.3.1.1 |  |  |  |  |  |  | 142(Q)L; 208(I)L;<br>297(N)Y | 276(N)S;<br>540(S)A |  |  | 103(I)T; 240(N)S;<br>1725(D)E/G;<br>2163(N)Y |
| A.D |  |  |  |  |  |  | 232(E)G*; 253(T)K*;<br>260dup>QEETLHST<br>TSEGYLSPSQVYTT<br>SG |  |  |  | 598(Y)H* |
| A.D.1 |  |  |  |  | 43(M)I |  | 142(L)S; 320 (T)A* |  |  |  | 143(N)D; 179(T)S/L;<br>1653(I)V; 1661(K)N |
| A.D.1.1 |  |  |  |  |  |  | 121(S)G; 273(Y)H |  |  | 2(T)I | 124(V)I; 234(I)V;<br>970(N)S; 1187(D)G;<br>1236(T)S |
| A.D.1.2 |  |  | 104(I)F |  | 25(K)R* | 51(H)Q | 129(T)I*; 174(S)N |  |  |  | 1789(S)F |
| A.D.1.3 | 9(I)L | 3(T)I |  |  |  |  | 100(S)I*; 225(V)A;<br>263(E)Q; 265(L)P;<br>270(S)P*; 277(S)P;<br>280(Y)H; 291(S)P;<br>297(Y)H; 310(L)P;<br>316(S)P*; 321(K)X;<br>753(D)G* |  |  |  | 755(D)G*; 2111(N)H |
| A.D.2 |  |  |  |  |  |  | 134(K)I; 262(E)K |  |  | 46(N)S | 598(H)Y*; 1731(D)G |
| A.D.2.1 |  |  |  | 69(T)A |  | 21(I)V | 67(H)N; 107(T)P;<br>120(P)T; 240(K)R;<br>280(Y)H; 287(E)D;<br>294(S)P; 303(V)I;<br>319(T)S |  |  |  | 144(G)E; 610(P)S;<br>1599(H)R |
| A.D.2.2 |  |  |  |  |  |  | 240(R)K; 243(I)S;<br>303(I)V |  |  |  | 1731(D)G; 1950(I)L |
| A.D.2.2.1 |  |  |  |  |  |  | 224(E)G; 265(L)I;<br>274(L)P |  |  |  | 171(L)P; 256(K)R |
| A.D.3 |  |  |  | 92(T)M* |  |  | 113(T)I; 131(V)D*;<br>178(N)G*; 258(H)Q*;<br>266(H)L* | 12(T)I |  | 79(T)A | 835(L)M |
| A.D.3.1 |  |  |  |  |  |  | 101(F)S; 151(R)H;<br>225(V)A; 230(P)T*;<br>250(S)F*; 263(E)K | 20(L)F;<br>276(S)N |  | 48(I)L |  |
| A.D.4 |  |  |  | 63(I)V/A | 47(L)Q |  | 57(A)T*; 206(P)Q;<br>310(L)P* |  |  |  | 1726(K)R |
| A.D.4.1 |  |  | 122(E)D |  |  |  | 90(Y)H; 111(S)F*;<br>283(S)P; 312(Q)X;<br>319(T)A | 123(K)Q;<br>384(I)T | 9(F)Y;<br>173(I)V | 1(T)M | 1617(V)I |
| A.D.5 |  |  |  |  |  |  | 206(P)Q; 303(V)A*;<br>320(T)A* | 122(T)A |  |  | 1723(S)G |
| A.D.5.1 |  |  | 216(Y)H |  |  |  | 57(A)V; 118(T)I*;<br>209(K)R; 319(T)I | 23(A)S* |  |  |  |
| A.D.5.2 |  |  |  | 69(T)I |  |  | 57(A)V; 209(K)R* | 103(A)T* |  | 26(C)Y* |  |
| A.D.5.3 |  | 3(T)A;<br>8(T)A |  |  |  |  | 189(I)T; 225(V)A;<br>243(I)T; 276(P)Q*;<br>280(Y)H; 284(G)D*;<br>314(L)P* | 15(L)F;<br>419(K)E | 52(K)R;<br>173(I)V* | 27(R)K;<br>64(D)E | 114(R)K; 179(T)A;<br>335(V)I; 1051(N)S |

**Table 2.**
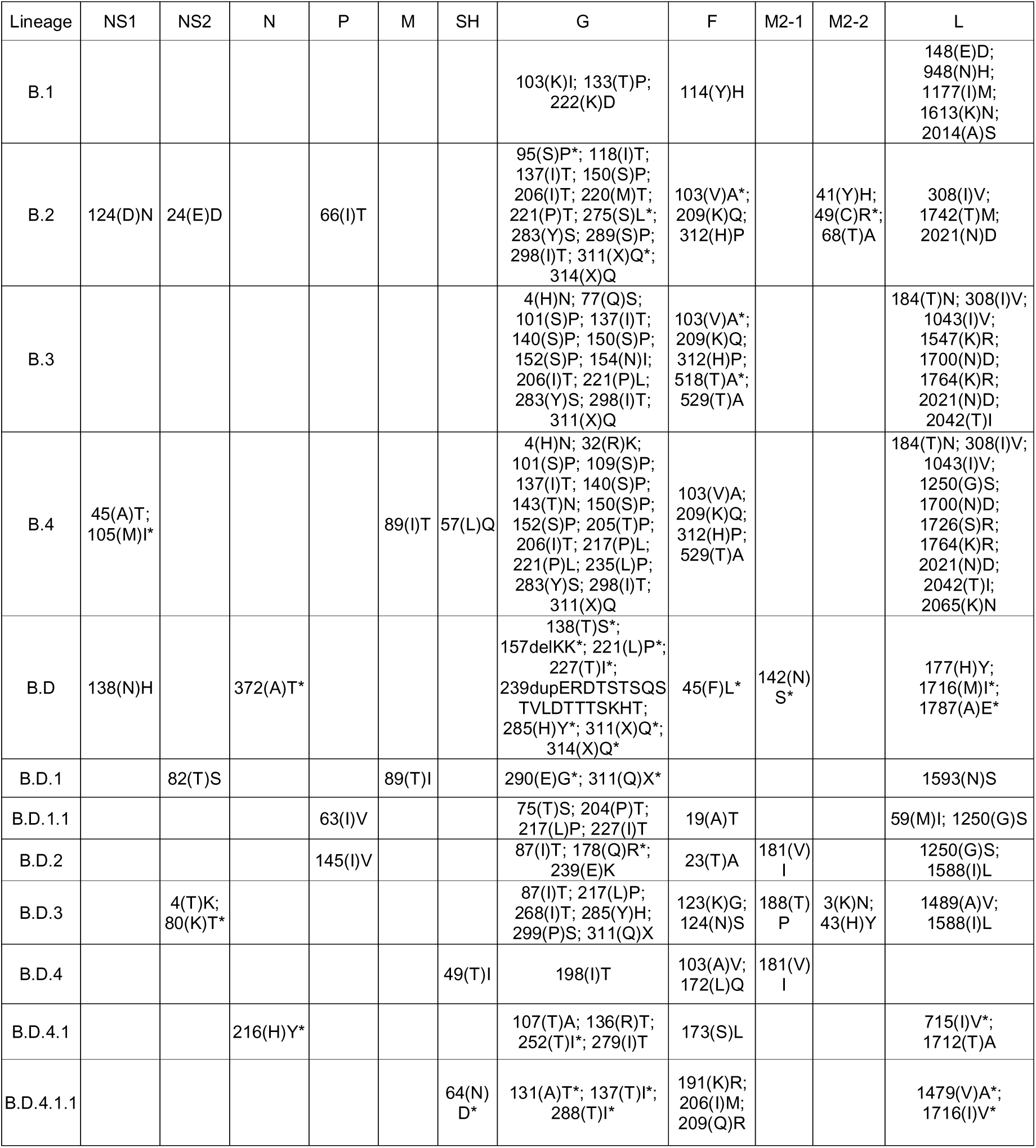
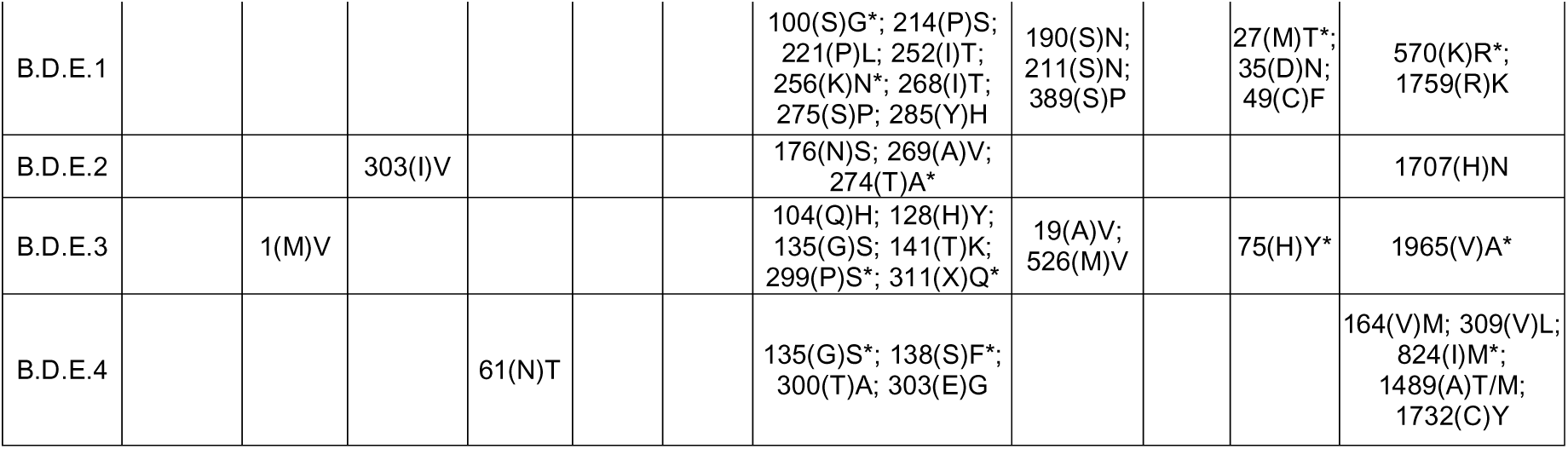
Amino acids defining lineages in HRSV-B. The lineage-defining amino acids within each viral protein are detailed. This description provides the location of the amino acid using the genome coordinates HRSV/B/AUS/VIC-RCH056/2019 (lineage B.D.4.1.1, NCBI Virus accession number: OP975389.1, GISAID EpiRSV accession number: EPI_ISL_1653999), followed by the amino acid found in the parental lineage in parentheses. Finally, the amino acid substitution that drives it to become a lineage is specified. Amino acid changes detected in not all but ≥90% of the lineage sequences are indicated with *. The letter X is used to represent an amino acid deletion.

### The use of G sequences with the HRSV lineage classification system

The main challenge for global expansion of HRSV genomics is the absence of a cost-effective, globally standardized and validated methodology for sequencing, in contrast to SARS-CoV-2 or influenza virus (36,37). In addition, limited funding and/or infrastructure causes some laboratories to prefer sequencing the G gene only (38–40). While we recommend the use of complete genomes for HRSV lineage assignment, partial genomes covering the G and/or F genes can be used since they reproduce the topology of the HRSV tree (16,18). As previous reports showed the decreased phylogenetic signal, we do not recommend the use of smaller G gene regions such as the second hypervariable region (250-nt length at the 3’ gene end, Figure 1) that was used historically for molecular epidemiology (16).

Minimal misclassification (1.2% error) was found in HRSV-A and none in HRSV-B when we explored the reproducibility of the classification using the G gene only, retaining strong support for lineage-defining nodes (Supplementary Material). However, the use of G gene limits the detection of emergent lineages and precludes monitoring of potential immune evasion in the F protein.

### Prospective HRSV lineage assignment and definition

The assignment of sequences to the existing lineages can be automated using online tools such as NextClade (https://clades.nextstrain.org/) (41), ReSVidex (https://cacciabue.shinyapps.io/resvidex/), INSaFLU (https://insaflu.insa.pt) (42) or UShER (https://usher.bio/) (43). However, for a classical approach to define the lineage of query sequences, we encourage users to follow our guidelines described in the Supplementary Material and available on GitHub (https://github.com/orgs/rsv-lineages/repositories).

We anticipate new lineages of HRSV-A/B will continue to emerge, and we envision expanding our proposed nomenclature to incorporate new lineages. Detection and definition of a new lineage should adhere to the considerations outlined in this study and described in our operative guidelines (Supplementary Material).

It is important to stress that assigning the lineage of a sequence does not require the completeness of the genome or the absence of nucleotide ambiguities, it only requires supported association in a phylogenetic tree. In this sense, interpretation of results when complete genomes are not used should consider the intrinsic limitations. However, to define a new lineage, complete genomes and the absence of ambiguities are mandatory due to the amino acid characterization as part of the definition. We encourage the general community to report detection of new HRSV lineages at the RGCC GitHub page as an ‘issue’ within the corresponding repository for HRSV-A/B. The RGCC study group will evaluate the newly proposed lineage, and if confirm, reference alignments will be updated.

### Molecular epidemiology of HRSV with the proposed classification

Using our proposal, we described the HRSV molecular epidemiology including all available genomes, even those previously discarded due to reasons such as the presence of nucleotide ambiguities. Consequently, we analyzed the seasonality of lineages using a dataset comprising 2,277 HRSV-A and 2,058 HRSV-B genomes, revealing notable co-circulation and lineage replacement over time (Figure 3C, Figure 4C). Within HRSV-A, A.1 and A.2 lineages are currently extinct, with the last detected sequences collected in 1995, and 2015, respectively. Since 2011, A.D and nested lineages continue to circulate. Descendant lineages within A.D (A.D.2.2 and A.D.4) were first detected in 2013, highlighting the rapid divergence of the HRSV-A viruses with the duplication in G gene. Within HRSV-B, the circulation of lineages B.1, B.2, B.3, and B.4 exhibited a strong lineage replacement (Figure 4C). Although the first reported detection of viruses with a 60-nt duplication in the G gene (B.D lineage) traces back to 1999, complete genomes only became available from 2005 onwards (8). By 2009, only genomes from B.D and nested lineages were detected, and from 2017 onwards, only B.D.4 and nested lineages have been observed.

Post-COVID-19 pandemic, a higher number of HRSV-A than HRSV-B lineages were detected, especially from A.D.1, A.D.3-A.D.5 divergent branches, while A.D.2 exhibited sporadically low detections. Post-pandemic HRSV-B viruses primarily classified within lineages B.D.4.1 and its divergent branches, B.D.4.1.1 and descendants B.D.E.1 – B.D.E.5. While this is an overall analysis about post-pandemic HRSV spread, further analysis is needed to understand global transmission, local evolution, and detect geographically limited lineages due to SARS-CoV-2 control measures. Pre-pandemic lineages may not have been described due to weak genomic surveillance. However, if pre- or post-pandemic HRSV genomes are shared publicly in the future, our classification will allow updates via RGCC GitHub when a new lineage is discovered.

With the current data, we observed that some lineages were only post-pandemic - A.D.3.1, A.D.5.2, and A.D.5.3 in HRSV-A, and B.D.E.1 and B.D.E.3 in HRSV-B. However, most of the lineage-defining amino acids in the new lineages were found pre-pandemic. For example, A.D.5.2 acquired lineage status when the substitution C26Y in M2-2 appeared, while the other four substitutions were already present in the closest genome of the parental lineage from 2019 (GenBank accession number MZ515825). It is important to mention the detection of the new post-pandemic lineages does not contradict recent studies stating the absence of new post-pandemic ‘genotypes’ as they were based on previous classification systems (44–47). In addition, the possibility that these new post-pandemic lineages have been circulating since before the pandemic depends on the deposition of genomes.

### Final remarks and conclusion

A consensus classification of HRSV below the subgroup level has been a challenge for multiple decades. Collaboratively, the HRSV molecular evolution research community, along with experts in the evolution of other respiratory viruses, have worked toward establishing a unified global classification system. This proposal categorizes HRSV-A/B sequences into lineages based on phylogenetic associations and amino acid markers, relying on complete genomes. While defining a new lineage requires the complete genome sequence, partial or low-quality genomes can be assigned to the existing lineages, emphasizing the robustness of this system. We developed standard guidelines for the lineage definition, lineage assignment, and created online resources for updates, ensuring long-term utility.

Defining a viral category below species through a phylogenetic-based classification is challenging. The system must exhibit reproducibility, balance complexity, and be updatable to capture the level of heterogeneity useful for viral surveillance. While different proposals may emerge, they may not succeed in their ability to address the specific questions for a given virus. Illustratively, three SARS-CoV-2 phylogenetic-classification were proposed by Nextstrain, GISAID, and Pango (48,49). However, Pango classification proved to be instrumental to monitor a rapidly evolving virus within a constrained temporal window and dealing with the meticulous detection of emergent mutations through unparalleled global genomic surveillance efforts.

HRSV is not an emerging virus; it generates annual outbreaks with co-circulation and replacement in the prevalence of its antigenic subgroups. Though few, there are RSV genomes collected from clinical samples collected >50 years ago. The largest increase in the number of genomes occurred in the last two years, and we anticipate this trend continuing. In 2023, recombinant F protein vaccines were approved for the first time. As their implementation progresses, we will learn how this affects viral evolution. We hope this unification proposal for the phylogenetic classification of HRSV will facilitate spatiotemporal comparative lineage surveillance and detection of emerging lineages. A limitation of our definition is the uncertainty regarding the impact of individual amino acid substitutions on lineages. We do not know a priori whether a simple amino acid substitution may have a significant epidemiological impact. Hence, whole genome surveillance and the study of the lineage-phenotype association will be essential, as observed in genetic and antigenic characterization in influenza to estimate the effectiveness of immunization (50). Other knowledge gaps include, but are not limited to, analyzing the association between lineages and the severity of HRSV disease, as well as associations of particular lineages with demographic characteristics of individuals.

## Data Availability

Viral genomes used in this study are openly available in the public databases NCBI GenBank (https://www.ncbi.nlm.nih.gov/genbank/) and GISAID EpiRSV (https://gisaid.org/).

https://github.com/rsv-lineages/Classification_proposal

## Acknowledgments

Authorships between the second and last were listed alphabetically by name. We acknowledge the authors who have shared HRSV genomes on the public databases NCBI GenBank, ENA, DDBJ, and GISAID EpiRSV. We express our gratitude for the valuable comments provided by researchers and public health scientists during the initial stages of the RSV Genotyping Consensus Consortium’s work.

## Disclaimers

RAN consults for Moderna on matter in virus evolution. NW has received grant funding from the Bill and Melinda Gates Foundation and Sanofi. The authors received no financial support for the research, authorship, or publication of this article.

## Notes

### Author Declarations

Viral genomes used in this study were openly available in the public databases NCBI GenBank (https://www.ncbi.nlm.nih.gov/genbank/) and GISAID EpiRSV (https://gisaid.org/).

