## Supplementary Material for "The unified proposal for classification of human respiratory syncytial virus below the subgroup level"

The description of each supplementary section appears in the order of mention within the manuscript.

Phylogenetic analysis.

Maximum likelihood phylogenetic trees were constructed with IQ-TREE v2.2.0, using ModelFinder to find the best nucleotide substitution model (1,2). The reliability of sequence clusters was evaluated with SH-aLRT (1,000 replicates) and UFBoot2 (10,000 replicates) (3). Monophyletic clades defining lineages were considered when SH-aLRT value was ≥80% and UFBoot2 value was ≥90%. Phylogenetic trees were visualized with Figtree v1.4.4 and Auspice (4,5). Temporal signal was assessed with TempEst v1.5.3, and molecular-clock phylogenies were inferred with TreeTime (6,7).

Recombination events were assessed with both alignment-based and phylogenetic-based methods. RDP4 software was used to detect and characterize the recombination events within the sequence alignments using the RDP, GENECONV, Maximum Chi Square and 3SEQ methods with default settings (8). The TreeKnit software assessed recombination based on topological differences between trees by comparison of phylogenies inferred with the 5’ and 3’ ends of the alignments (4,500-nt each, excluding the G gene) (9). The resulting tanglegram, available at <https://github.com/rsv-lineages/Classification_proposal>, was visualized in Auspice. Recombination assessment, utilizing RDP4 software (alignment-based) and phylogenetic tree topology-based analysis, found no evidence of recombination among HRSV sequences. Consequently, no sequences were removed due to genetic recombination.

Defining the root and the final dataset of HRSV phylogenetic trees.

Two approaches were used to define the correct phylogenetic tree root: a) the utilization of an outgroup, a conventional method for inferring the tree root using sequences known to be evolutionarily distant, and b) phylodynamic analysis, integrating temporal and phylogenetic patterns in virus evolution.

In the first approach, five HRSV-B genomes with the earliest collection dates were aligned with the HRSV-A dataset, and vice versa (Supplementary Figure 3A, 3C). As anticipated, the five sequences from the alternative subgroup formed a distantly related clade, serving as the outgroup for rooting each tree. The second approach, without outgroups, involved reconstructing dated phylogenetic trees for each subgroup dataset, and the phylogenetic root automatically inferred by incorporating temporal information (Supplementary Figure 3B, Figure 3D). Both approaches consistently identified the same root for each subgroup cluster.

Comprehensive characterization of the datasets was achieved through phylodynamic analysis. The most recent common ancestor (MRCA) was dated to 1951 for HRSV-A and 1965 for HRSV-B (Supplementary Figure 3E-F). Global evolutionary rates were estimated to 7.964x10^-4^ substitutions/site/year (r^2^=0.96) for HRSV-A and 6.933x10^-4^ substitutions/site/year (r^2^=0.98) for HRSV-B, aligning with previous reports (10–12).

Outlier sequences in root-to-tip plots evaluating the temporal signal may be unreliable due to, for example, sequencing errors and/or inaccurate metadata. We identified 58 outlier sequences for HRSV-A and 2 for HRSV-B that were excluded from rates estimation and lineage designation. Following the exclusion of outliers from the root-to-tip analysis, the final dataset considered for lineage designation comprised 1,480 HRSV-A genomes and 1,385 HRSV-B genomes.

The HRSV lineage definition.

Results of the phylogenetic clustering using from 1 to 10 amino acid as thresholds in an automated manner are available at <https://github.com/rsv-lineages/Classification_proposal>.

The use of G gene sequences with the HRSV lineage classification system.

Reproducibility of the classification with the G gene sequences was assessed. The phylogenetic trees obtained are available at <https://github.com/rsv-lineages/Classification_proposal>.

HRSV lineage assignment of query sequences.

The assignment of sequences to the existing lineages can be automated using online tools such as NextClade (<https://clades.nextstrain.org/>) (13), ReSVidex (<https://cacciabue.shinyapps.io/resvidex/>), INSaFLU ([https://insaflu.insa.pt](https://urldefense.com/v3/__https:/insaflu.insa.pt/__;!!K-Hz7m0Vt54!gdGL4_tqYti7Zfk3bbG1jRTB8P7zUnC1hcjmPEgSvKt0_Co8Z88EXzjXTXBInezTN25k5BjGzmqkI0tLFBCGMZcmpYy8$)) (14,15) or UShER (<https://usher.bio/>) (16). However, for the classical approach to define the lineage of query sequences we encourage users to follow the guidelines described below:

1. Perform an alignment (for instance, with MAFFT, online version (17): <https://mafft.cbrc.jp/alignment/server/>) with the query sequences and the reference sequences of the same HRSV subgroup available in the GitHub (<https://github.com/rsv-lineages/lineage-designation-A> and <https://github.com/rsv-lineages/lineage-designation-B>).
2. Visually verify that the alignment covers the entire genomic region from the first codon of NS1 gene to the codon of the L gene. If the target sequences are longer than the reference alignments, trim the ends accordingly. If the target sequences are shorter, no trimming is necessary.
3. Visually evaluate the alignment, especially around the G gene region, to detect and correct alignment artifacts.
4. Infer a maximum likelihood phylogenetic tree with software such as IQ-TREE enabling the selection of the nucleotide substitution model with ModelFinder and assessing node support with UFBoot2 and SH-alrt. Phylogenetic analysis can be run online (<http://iqtree.cibiv.univie.ac.at/>) (18), or locally using the command line as follows (version IQ-TREE v2.2.0):

*iqtree2 -s <path_to_sequences> --alrt 1000 -B 1000*

5. Visualize the phylogenetic tree with software such as FigTree. Find the lineage of the query sequences by associating them with the reference sequences. Evaluate whether the query sequences form a monophyletic clade with statistical support (≥90% for UFBoot2 and ≥80% for SH-alrt).

If, for example, G gene sequences will be used, it is recommended to trim the reference alignment from GitHub to the longest G open reading frame and verify the alignment including the query sequences mainly around the duplication region. Interpretation of results when complete genomes are not used should take into consideration the intrinsic limitations.

Prospective identification of new HRSV lineages.
We anticipate that new lineages of HRSV-A and HRSV-B will continue to emerge in future and envision our proposed nomenclature being expanded to incorporate new lineages. The detection and definition of a new lineage comprise the use of complete genomes that should adhere to the considerations outlined in this study on the phylogenetic and amino acid criteria. The recommended procedure to define a new lineage is described below:

1. Perform an alignment (for instance, with MAFFT, online version (17): <https://mafft.cbrc.jp/alignment/server/>) of the complete genomes of the potential new lineage with the reference alignment available on GitHub (<https://github.com/rsv-lineages/lineage-designation-A> and <https://github.com/rsv-lineages/lineage-designation-B>).
2. Ensure that complete genomes cover information from the first codon of the first gene (NS1) to the last codon of the last gene (L) without the presence of ambiguous nucleotides. Trim the alignment if necessary to meet the defined criteria for complete genomes. Conversely, sequences can include missing data only in the first or last 50 nucleotides of the alignment. Visually inspect the alignment to identify and correct bioinformatics artifacts, particularly in the G gene region.
3. Infer a maximum likelihood phylogenetic tree with IQTREE, enabling the selection of the nucleotide substitution model using ModelFinder, and assessing statistical support for nodes with UFBoot2 and SH-alrt. Phylogenetic analysis can be run online (<http://iqtree.cibiv.univie.ac.at/>) (18), or by the command line as follows (IQ-TREE v2.2.0):

*iqtree2 -s <path_to_sequences> --alrt 1000 -B 1000*

4. Evaluate the phylogenetic tree (in IQTREE v2.2.0, the file with a “.treefile” extension) to confirm that the potential new lineage meets the phylogenetic lineage criteria. This entails a monophyletic clade with at least 10 sequences of interest, supported by statistical values ≥90% for UFBoot2 and ≥80% for SH-alrt. The potential lineage should not contain sequences from any other lineage.
5. Conduct a comparative analysis of the protein sequences of the potential lineage and its parental lineage to confirm the amino acid lineage criteria. Subset the complete genomes alignment into individual open reading frames and translate the nucleotides using the universal genetic code. Identify among all the viral proteins at least 5 amino acid substitutions differentiating the potential lineage from the parental one. Those amino acids should present in more than 90% of the new lineage sequences.
6. If the potential lineage meets both the phylogenetic and amino acid substitution requirements, it can be named following the lineage nomenclature, including the suffix "x" to denote its potential status. For example, A.D.1.3.1pot for a descendant of A.D.1.3 and B.D.E.2.1x for a descendant of B.D.E.2. Another example based on the current state is A.D.E.1x for a descendant of A.D.2.2.1, where E is the alias for 2.2.1.
7. Share the proposal of the new lineage on the RGCC GitHub page as an 'issue' within the corresponding repository for HRSV-A (https://github.com/rsv-lineages/lineage-designation-A) or HRSV-B (https://github.com/rsv-lineages/lineage-designation-B). The RGCC study group will evaluate the new proposed lineage, and if accepted, the reference alignments will be updated.
