## Supplementary Figure 1 for "The unified proposal for classification of human respiratory syncytial virus below the subgroup level"

**Supplementary Figure 1. Dataset curation for the HRSV classification definition.** For each of the filtration step during the dataset curation the number of remaining HRSV-A and HRSV-B genomes is detailed.

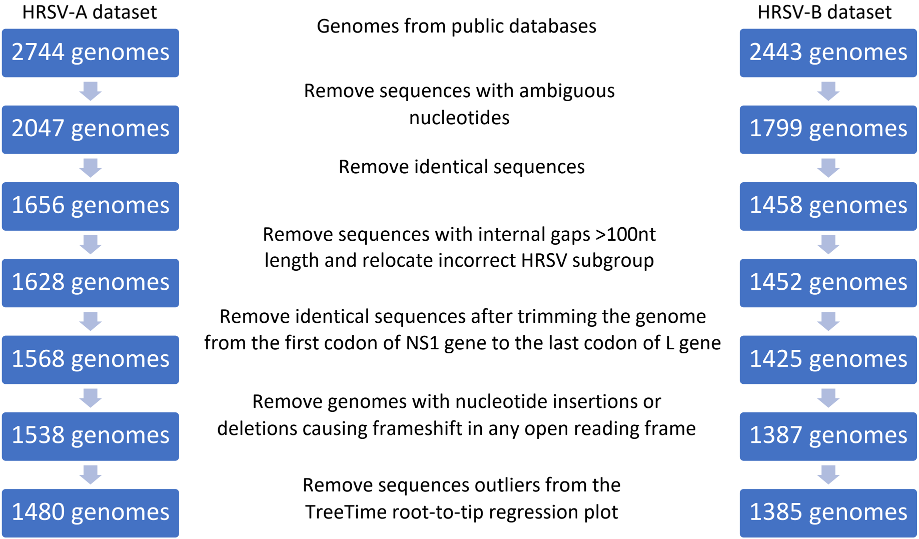
