## Supplementary Figure 2 for "The unified proposal for classification of human respiratory syncytial virus below the subgroup level"

Supplementary Figure 2. Characterization of the genomes used in the classification.
The number of HRSV genomes per year and the country of sample collection are reported. Countries are grouped into geographic regions highlighted in gray scale. The heat map highlights the countries representation by collection year.

| RSV genomes | Year | Argentina | Brazil | Canada | Nicaragua | USA | Cote d'Ivoire | Egypt | Kenya | Morocco | South Africa | Uganda | Cambodia | China | India | Israel | Japan | Jordan | Lebanon | Philippines | Russia | Thailand | Vietnam | Australia | New Zealand | Austria | Belgium | Bosnia and Herzegovina | France | Germany | Macedonia | Netherlands | Spain | Switzerland | United Kingdom |
|---|---|---|---|---|---|---|---|---|---|---|---|---|---|---|---|---|---|---|---|---|---|---|---|---|---|---|---|---|---|---|---|---|---|---|---|
| 1 | 1956 | | | | | 1 | | | | | | | | | | | | | | | | | | | | | | | | | | | | | |
| 1 | 1979 | | | | | 1 | | | | | | | | | | | | | | | | | | | | | | | | | | | | | |
| 8 | 1980 | | | | | 8 | | | | | | | | | | | | | | | | | | | | | | | | | | | | | |
| 2 | 1981 | | | | | 2 | | | | | | | | | | | | | | | | | | | | | | | | | | | | | |
| 9 | 1982 | | | | | 9 | | | | | | | | | | | | | | | | | | | | | | | | | | | | | |
| 1 | 1983 | | | | | 1 | | | | | | | | | | | | | | | | | | | | | | | | | | | | | |
| 4 | 1984 | | | | | 4 | | | | | | | | | | | | | | | | | | | | | | | | | | | | | |
| 5 | 1985 | | | | | 5 | | | | | | | | | | | | | | | | | | | | | | | | | | | | | |
| 3 | 1986 | | | | | 3 | | | | | | | | | | | | | | | | | | | | | | | | | | | | | |
| 5 | 1987 | | | | | 5 | | | | | | | | | | | | | | | | | | | | | | | | | | | | | |
| 1 | 1988 | | | | | 1 | | | | | | | | | | | | | | | | | | | | | | | | | | | | | |
| 2 | 1989 | | | | | 2 | | | | | | | | | | | | | | | | | | | | | | | | | | | | | |
| 3 | 1990 | | | | | 3 | | | | | | | | | | | | | | | | | | | | | | | | | | | | | |
| 4 | 1991 | | | | | 4 | | | | | | | | | | | | | | | | | | | | | | | | | | | | | |
| 2 | 1992 | | | | | 2 | | | | | | | | | | | | | | | | | | | | | | | | | | | | | |
| 2 | 1993 | | | | | 2 | | | | | | | | | | | | | | | | | | | | | | | | | | | | | |
| 13 | 1994 | | | | | 13 | | | | | | | | | | | | | | | | | | | | | | | | | | | | | |
| 13 | 1995 | | | 4 | | 9 | | | | | | | | | | | | | | | | | | | | | | | | | | | | | |
| 1 | 1996 | | | | | 1 | | | | | | | | | | | | | | | | | | | | | | | | | | | | | |
| 20 | 2001 | | | | | 20 | | | | | | | | | | | | | | | | | | | | | | | | | | | | | |
| 9 | 2002 | | | | | 9 | | | | | | | | | | | | | | | | | | | | | | | | | | | | | |
| 15 | 2003 | | | | | 15 | | | | | | | | | | | | | | | | | | | | | | | | | | | | | |
| 5 | 2004 | | | | | 5 | | | | | | | | | | | | | | | | | | | | | | | | | | | | | |
| 4 | 2005 | | | | | 4 | | | | | | | | | | | | | | | | | | | | | | | | | | | | | |
| 7 | 2007 | | | | | | | | 7 | | | | | | | | | | | | | | | | | | | | | | | | | | |
| 50 | 2008 | | 43 | | | 1 | | | 1 | | | | | | | | | | | | | | | | | | | | | | | | | | 5 |
| 55 | 2009 | | 38 | | | 1 | | | 1 | | | | | 1 | | | | | | | | | | | | | | | | | | | | | 14 |
| 99 | 2010 | | 31 | | | | | | 51 | | | | | | | | | 3 | | | | | | | | | | | | | | | | | 14 |
| 22 | 2011 | | | | | | | | 2 | | | | | | | | | 12 | | | | 1 | | | | | | | | | | | | | 7 |
| 114 | 2012 | | | | | 3 | | | 67 | 7 | | | | 1 | | | | 2 | | 7 | | 1 | | | | | | | | | | | | | 26 |
| 58 | 2013 | | | | 1 | 4 | | | 16 | | | | | 1 | | | | 10 | | 6 | | 2 | | | | | | | | | | | | | 18 |
| 112 | 2014 | 53 | | | | 2 | | | 17 | 13 | | | | 4 | | | | | 1 | 4 | | 2 | | | | | | | | | | | | | 16 |
| 141 | 2015 | 56 | | | 27 | 14 | | | 17 | 13 | | | | | | | | | | 2 | | | | 2 | | | | | | 2 | | | | | 8 |
| 161 | 2016 | 48 | | | 24 | 13 | | | 4 | 48 | | | | | | | 1 | | | 2 | | 2 | | 1 | | | | | | | | | | | 18 |
| 134 | 2017 | 47 | | | | 1 | 6 | | | 40 | | | | 2 | 1 | | | | | | | 2 | 2 | 2 | | | | | | | | 22 | 3 | | 6 |
| 164 | 2018 | 17 | | | | 1 | 2 | | | 5 | | | | 4 | | | | | | | 2 | | 2 | 4 | | | | | | | | 48 | 10 | | 69 |
| 215 | 2019 | 5 | 1 | | | 9 | 4 | 2 | | 15 | | | | | | | | | | 24 | 9 | | | 4 | | 2 | | | | | | 53 | 31 | 1 | 55 |
| 196 | 2020 | | | | | 2 | | 3 | | | | | | 17 | | | | | | 4 | 4 | | 2 | 140 | | 1 | | | | | | 2 | 11 | | 10 |
| 741 | 2021 | 8 | 1 | | | 6 | | | | | 58 | 1 | 1 | 19 | | 2 | | | | | 1 | | | 182 | 390 | 1 | | | 8 | | | | | | 63 |
| 463 | 2022 | | 1 | | | 72 | | | | | 33 | | | | | | | | | | | | | 250 | | 3 | 1 | 1 | | | 4 | | | | 98 |
