## Supplementary Figure 3 for "The unified proposal for classification of human respiratory syncytial virus below the subgroup level"

Supplementary Figure 3. Rooting the HRSV phylogenetic trees.
(A-B) HRSV-A maximum-likelihood (ML) phylogenetic tree rooted using an outgroup of HRSV-B sequences highlighted in blue is shown as well as the tree rooted by phylodynamic analysis. (C-D) Similarly in HRSV-B, the ML phylogenetic tree rooted using an outgroup of HRSV-A highlighted in blue and the tree rooted by phylodynamic analysis are shown. The branch indicating the common ancestor of the HRSV-A or HRSV-B sequences is indicated in red. (E-F) The genetic distances from the root to the tips of the ML tree are plotted against the year of collection. Root-to-tip mutation counts vs. sample collection date for HRSV-A and HRSV-B. Regression was plotted using to the best fitting root which minimizes the sum of the squared residuals from the regression line. The x-intercept of the regression line represents the estimated time of the most recent common ancestor of the dataset in the tree, while the gradient estimates the evolutionary rate. Outliers excluded from rate estimation are indicated.

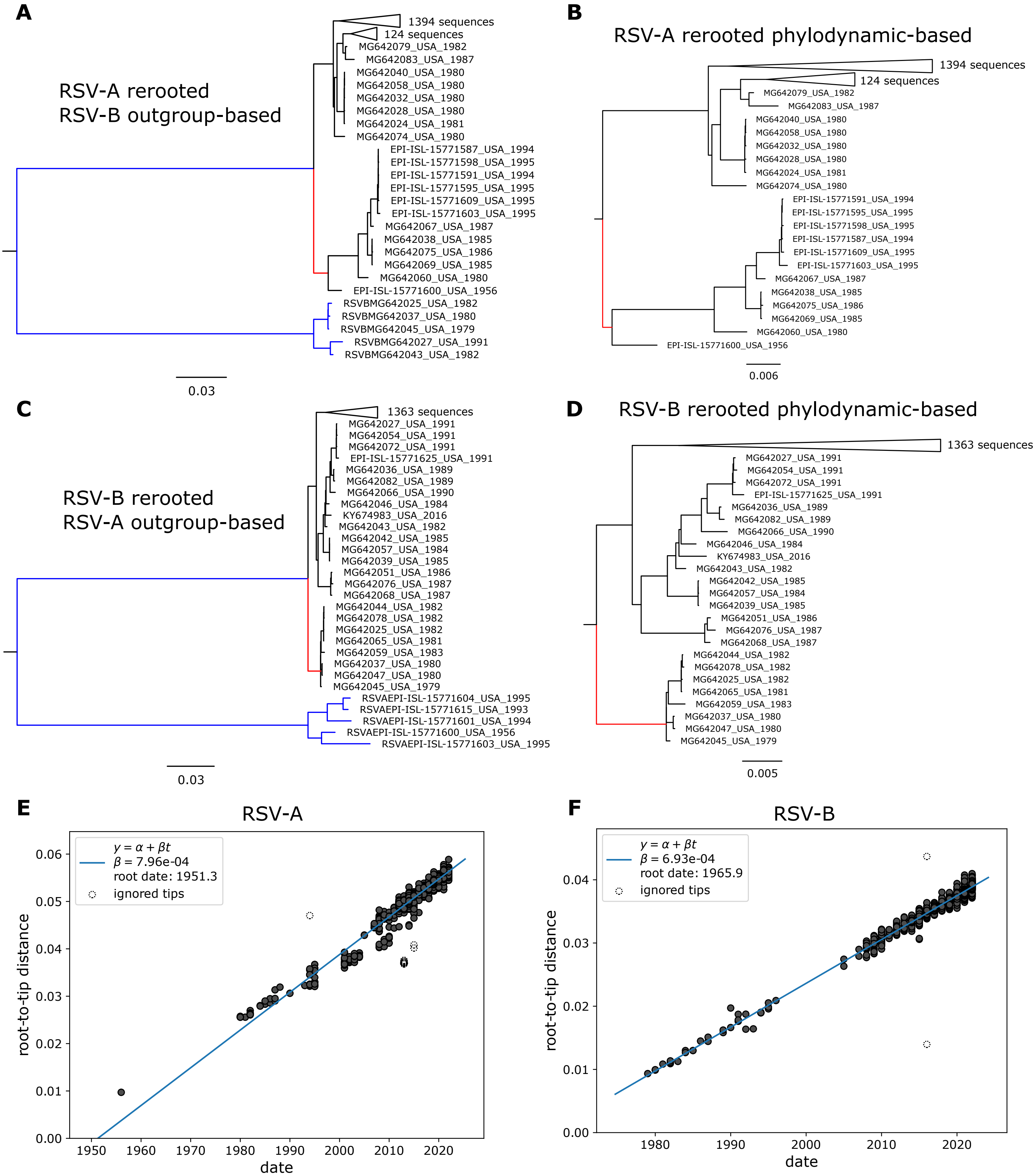
