## Supplementary Figure 4 for "The unified proposal for classification of human respiratory syncytial virus below the subgroup level"

Supplementary Figure 4. Allocation of HRSV-A and HRSV-B without the duplication in the G gene.
Genomes lacking the duplication of G gene in the dataset for the classification definition that were found within the clade containing the duplication of G are denoted with a red circle in the HRSV-A or HRSV-B maximum likelihood tree. The clade of A.D. or B.D. and nested lineages in HRSV-A or HRSV-B, respectively is denoted with a blue dashed line box.

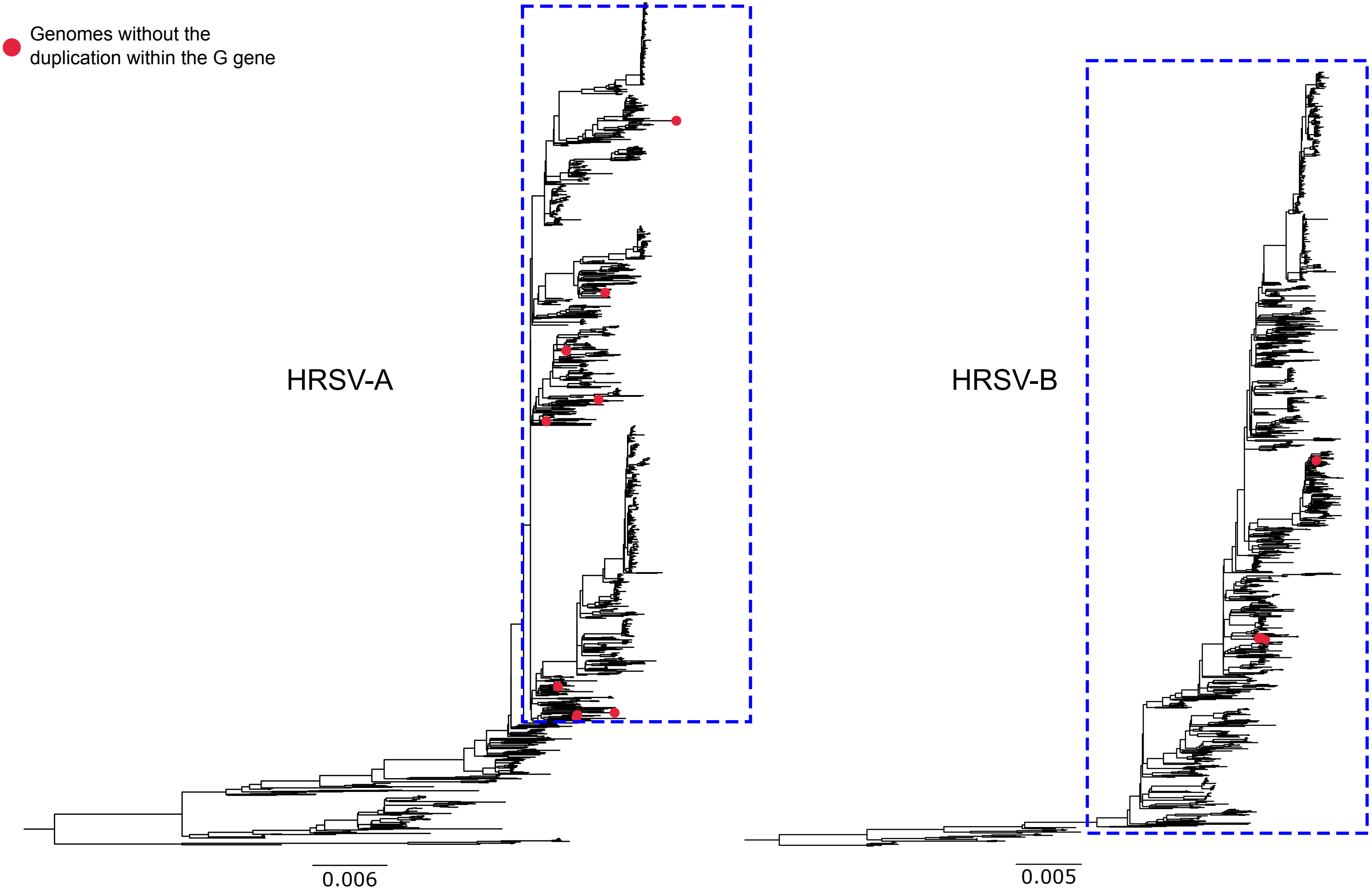
